# First dose mRNA vaccination is sufficient to reactivate immunological memory to SARS-CoV-2 in ex COVID-19 subjects

**DOI:** 10.1101/2021.03.05.21252590

**Authors:** Alessio Mazzoni, Nicoletta Di Lauria, Laura Maggi, Lorenzo Salvati, Anna Vanni, Manuela Capone, Giulia Lamacchia, Elisabetta Mantengoli, Michele Spinicci, Lorenzo Zammarchi, Seble Tekle Kiros, Maria Grazia Colao, Paola Parronchi, Cristina Scaletti, Lucia Turco, Francesco Liotta, Gian Maria Rossolini, Lorenzo Cosmi, Alessandro Bartoloni, Francesco Annunziato, for the COVID-19 Research Group

**Affiliations:** Department of Experimental and Clinical Medicine, University of Florence, Florence, Italy; Infectious and Tropical Diseases Unit, Careggi University Hospital, Florence, Italy; Microbiology and Virology Unit, Careggi University Hospital, Florence, Italy; Immunology and Cell Therapy Unit, Careggi University Hospital, Florence, Italy; Health Director, Careggi University Hospital, Florence, Italy; Flow cytometry diagnostic center and immunotherapy, Careggi University Hospital, Florence, Italy

## Abstract

Characterizing the adaptive immune response to COVID-19 vaccination in individuals who recovered from SARS-CoV-2 infection may define current and future clinical practice. To determine the effect of two doses BNT162b2 mRNA COVID-19 vaccination schedule in individuals who recovered from COVID-19 (ex COVID-19) compared to naïve subjects we evaluated SARS-CoV-2 spike-specific T and B cell responses, as well as specific IgG, IgM and neutralizing antibodies titres in 22 individuals who received BNT162b2 mRNA COVID-19 vaccine, 11 of which had a previous history of SARS-CoV-2 infection. Evaluations were performed before vaccination and then weekly until 7 days post second injection. Data obtained clearly showed that one vaccine dose is sufficient to increase both cellular and humoral immune response in ex COVID-19 subjects without any additional improvement after the second dose. On the contrary, the second dose is mandatory in naïve ones to further enhance the response. These results question whether a second vaccine jab in ex COVID-19 subjects is required and indicate that millions of vaccine doses may be redirected to naïve individuals, thus shortening the time to reach herd immunity.

## Introduction

As of February 26th 2021, over 112,6 million people have been diagnosed with COVID-19 worldwide, with more than 2,5 million confirmed deaths [1]. COVID-19 is associated with high transmission and, without effective treatments, rising numbers of cases of respiratory distress are threatening to overwhelm global health-care capacity [2-5]. Up to now, few COVID-19 vaccine based on mRNA (BNT162b2 and mRNA-1273) or adenovirus (ChAdOx1 nCoV-19) have been approved by international and national Medicines Agencies, making some difficulties in producing a number of vaccine doses adequate to meet current needs [6-9]. Approved COVID-19 mRNA vaccines require a first dose followed 3 to 4 weeks later by a second injection [6-7]. In many countries, vaccinations are carried out on all volunteer subjects regardless of their previous infection by SARS-CoV-2 [10]. This approach raises two main questions: a) is it necessary to vaccinate those who recovered from COVID-19? If confirmed, b) is it necessary to administer the second dose of vaccine as well?

In this manuscript, we evaluated SARS-CoV-2 spike-specific T and B cells response, as well as IgG, IgM and neutralizing antibodies specific titers in 22 individuals, 11 of which had a previous history of COVID-19 (ex COVID-19). The subjects were evaluated before receiving BNT162b2 mRNA COVID-19 vaccine and then weekly until 7 days post second injection, that was administered 21 days after the first one. Data obtained clearly showed that one vaccine dose is sufficient to increase both cellular and humoral immune response in ex COVID-19 subjects without any additional improvement after the second dose. On the contrary, the second dose is mandatory in naïve ones. These results question whether a second vaccine jab in ex COVID-19 subjects is indeed required and indicate that millions of vaccine doses may be redirected to naïve individuals.

## Results and Discussion

During the first two months of the vaccination campaign against COVID-19 made with BNT162b2 mRNA COVID-19 vaccine, we recruited 22 healthcare workers, 11 of which had a previous history of SARS-CoV-2 infection. The demographic and clinical characteristics of the 22 recruited subjects and time from recovery to vaccination are detailed in Supplementary Table 1. 22 subjects were enrolled and had a blood draw at basal time (time 0, before injection of the first dose), then at 7 days, 14 days, 21 days (before injection of the second dose), and 28 days after (Supplementary Fig.1). The 22 recruited subjects were evaluated at each time point for humoral response and the presence of SARS-CoV-2 spike-specific T and B cells.

As shown in Figure 1A, anti-nucleoprotein (N) IgG were present before vaccination only in ex COVID-19 subjects, confirming their previous exposure to SARS-CoV-2 (p<0.001). As expected, the levels of anti-N IgG remained stable over the study period in ex COVID-19 subjects and did not raise in naïve individuals (Fig.1A). Instead, anti-S IgM increased over time in naïve subjects, while exhibiting no substantial changes in ex COVID-19 individuals (Fig.1B). Pre-vaccination levels of anti-S IgG were present only in ex COVID-19 subjects (p<0.01). In naïve individuals, anti-S IgG increased progressively from 14 to 21 days following the first injection and further raised at day 28, one week after the second jab. On the contrary, in ex-COVID-19 subjects anti-S IgG massively increased already at day 7 following first injection, reaching the upper detection limit, and remained stable over time (Fig.1C). We also measured anti-RBD IgG, which showed a kinetic similar to total anti-S IgG in both naïve and ex-COVID-19 individuals (Fig.1D). Moreover, an interesting observation emerged from the determination of neutralizing antibody titers. These antibodies were already present in the sera of ex COVID-19 subjects but significantly increased after the first BNT162b2 mRNA vaccine dose and they stabilized over time even after the second jab (Figure 1E). On the other side, naïve subjects showed appreciable titers of neutralizing antibodies only one week after the administration of the second vaccine dose and, even in this case, did not reach the levels present in ex COVID-19 subjects’ sera (Figure 1E).

**Figure 1.**
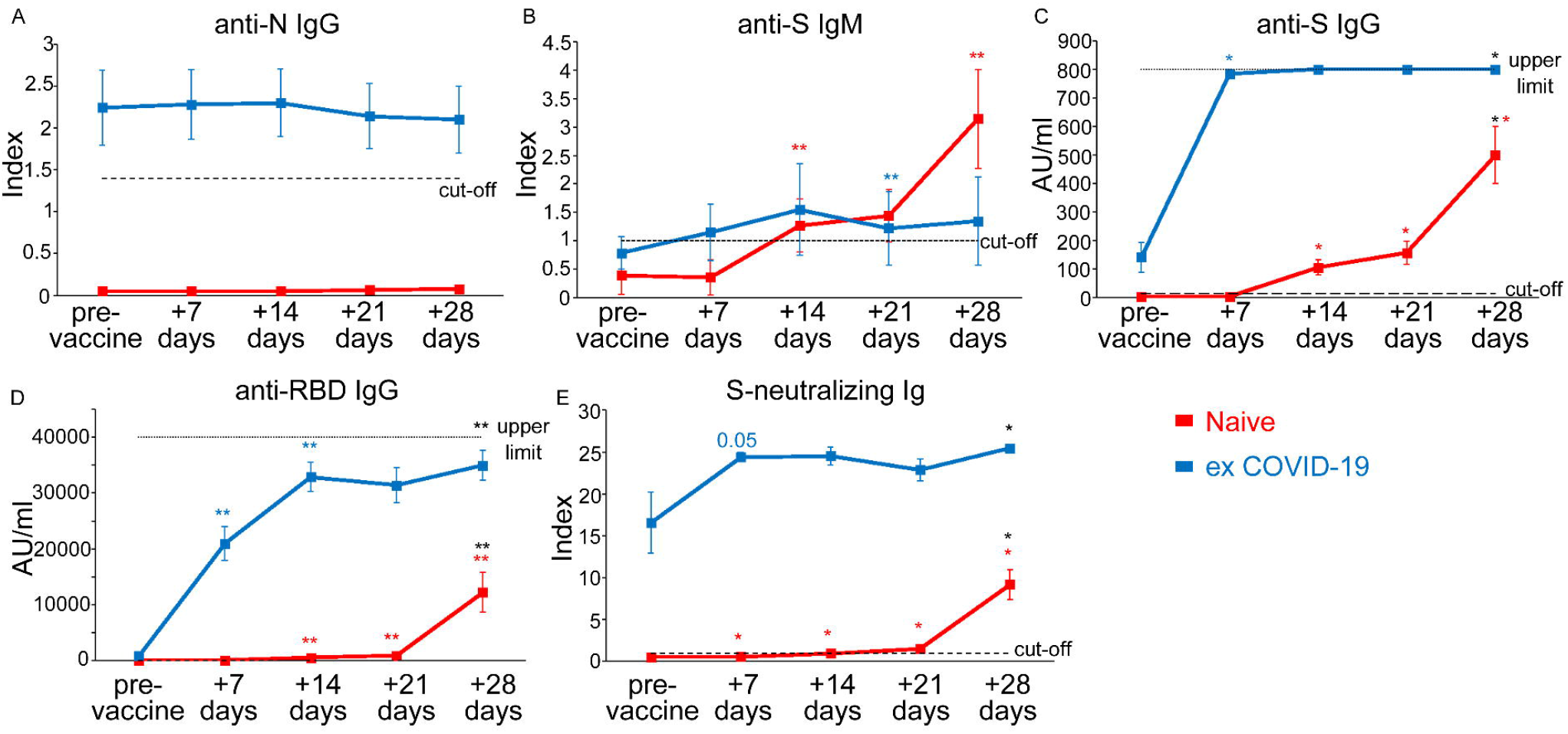
Evaluation of anti-SARS-CoV-2 serum antibody levels. Evaluation of anti-N IgG (A), anti-S IgM (B), anti-S IgG (C), anti-RBD IgG (D), S-neutralizing antibodies in naïve (red lines) and ex COVID-19 (blue lines) subjects before and after 7, 14, 21, 28 days of first vaccine administration. Data are represented as mean ±SE from 11 naïve and 11 ex COVID-19 subjects (A,B,D) or from 6 naïve and 6 ex COVID-19 subjects (C, E). Dashed lines represent cut-off values, dotted lines are upper detection limits. Blue and red asterisks refer to paired statistics within each study group compared to the previous time point in the kinetic. Black asterisks at day 28 represent paired statistic compared to pre-vaccine point. *= p<0.05; **= p<0.01; ***= p<0.001.

To further support the data obtained from the analysis of the serum levels of SARS-CoV-2 spike-specific antibodies, we evaluated by flow cytometry the frequency of B cells capable of recognizing SARS-CoV-2 spike protein (Figure 2A and Supplementary Figure 2). As shown in Figure 2B, ex COVID-19 subjects already had significantly higher frequencies of spike protein reactive B cells before vaccination as compared to naïve individuals (p<0.001). In ex COVID-19 subjects we observed a constant increase of the frequency of spike protein-reactive B cells up to 21 days, but then significantly declined after the second jab (Figure 2B). On the contrary, naïve individuals showed an appreciable increase of these cells, only one week after the second dose (Figure 2B). Of note, the frequency of spike-specific B cells at day 28 was significantly lower in naïve than in ex COVID-19 subjects (p<0.001). As shown in Figure 2C, D and E, SARS-CoV-2 spike protein-reactive B cells were predominantly CD27+ IgG+ cells, but CD27+ B cells of the IgM and IgA isotype could also be identified). Ex COVID-19 subjects had a kinetics of B cells expressing the three different immunoglobulin classes comparable to that described for total B cells reactive for the spike protein (Figure 2C, D, and E). Also in this case, in naïve subjects, an increase in the frequency of these cells was appreciated only after the administration of the second dose of vaccine ((Figure 2C, D, and E).

**Figure 2.**
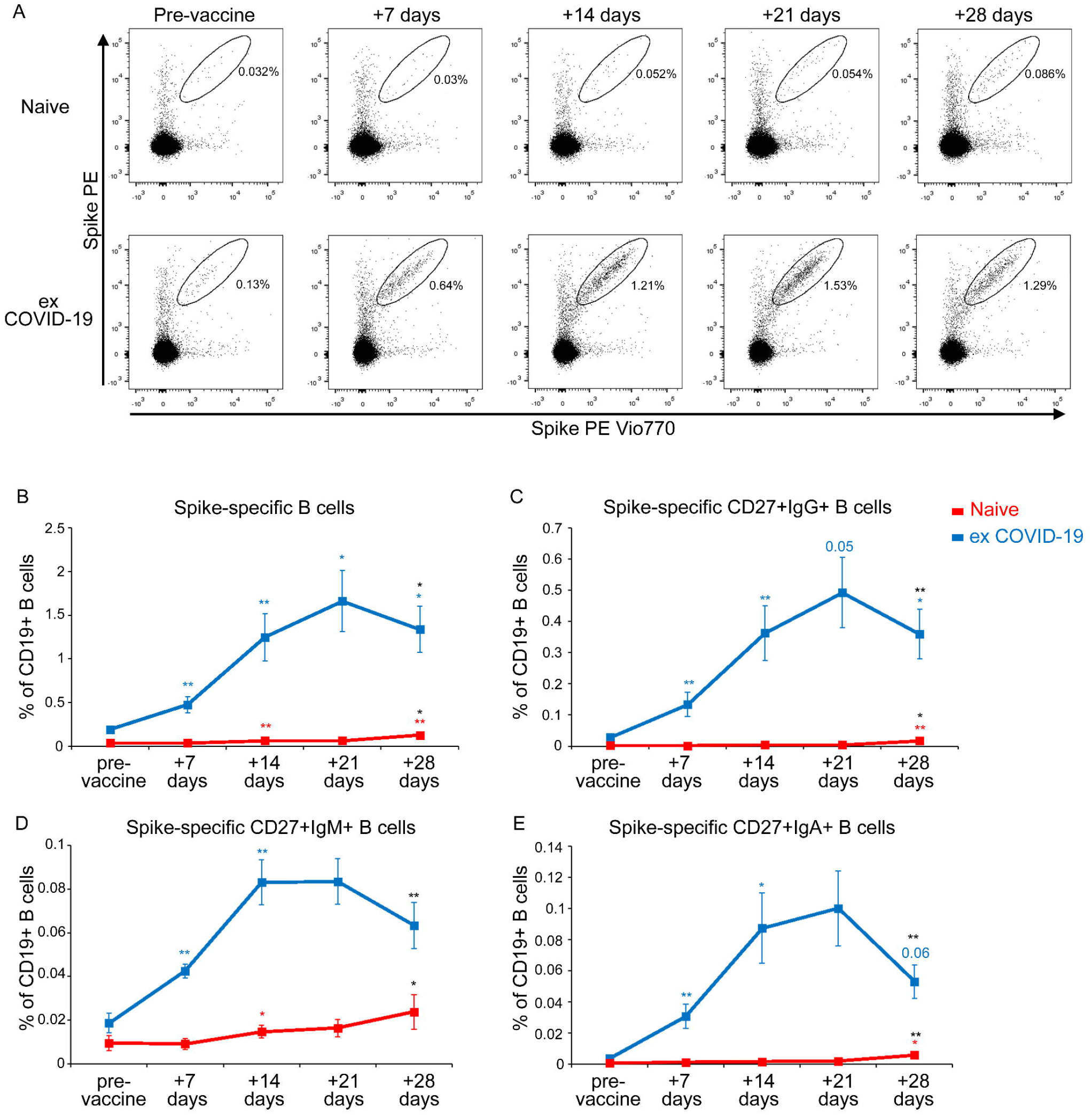
Detection of Spike-specific circulating B cells in naïve and ex COVID-19 vaccinated subjects. (A) Representative flow cytometric plots of Spike-specific B cells in one naïve (upper row) and one ex COVID-19 subject (lower row) before vaccination and after 7, 14, 21, 28 days from the first injection. Kinetic analysis of frequencies of total (B), CD27+ IgG+ (C), CD27+ IgM+ (D), CD27+ IgA+ (E) Spike-specific B cells in naïve (red lines) and ex COVID-19 (blue lines) subjects, before and after vaccination. Data are represented as mean ±SE from 11 naïve and 11 ex COVID-19 subjects. Blue and red asterisks refer to paired statistics within each study group compared to the previous time point in the kinetic. Black asterisks at day 28 represent paired statistic compared to pre-vaccine point. *= p<0.05; **= p<0.01; ***= p<0.001.

The data obtained from the study of B cells and from the evaluation of the serum levels of SARS-CoV-2 spike-specific antibodies, including those with neutralizing capability, showed that while the first administration of vaccine is sufficient in ex COVID-19 subjects to reactivate immunological memory, in naïve individuals the second BNT162b2 mRNA vaccine dose is mandatory. However, even a second jab does not allow naïve subjects, at least at day 28, to obtain serum Ig levels, in particular those with neutralizing capacity, as well as frequency of circulating spike reactive B cells, comparable to those present in ex COVID-19 subjects.

After evaluating B cell response, we also studied the antigen-specific CD4+ T cell response to SARS-CoV-2 spike protein peptides, monitoring CD154 expression and the production of IL-2, IFN-γ and TNF-α upon in vitro stimulation (Figure 3A and Supplementary Figure 3). Similarly to what we observed with B cells, SARS-CoV-2 spike-specific CD4+ T cells were present in the peripheral blood of ex COVID-19 subjects before vaccination, while not in naïve individuals (p<0.01) (Figure 3B). Moreover, in ex COVID-19 subjects we found a significant increase at day 7 after the administration of the first dose of BNT162b2 mRNA vaccine, in contrast to naïve individuals (Figure 3B). This observation is evident not only by evaluating the frequency of CD4+ T cells expressing CD154 and at least one of the three monitored cytokines (IL-2, IFN-γ and TNF-α) (Figure 3B), but also by evaluating the frequency of CD154-expressing T CD4+ cells in association with individual cytokines (Figure 3 C, D and E). Unlike what we found for B cell response, the frequency of spike-specific CD4+ T cells showed a trend towards a reduction in ex COVID-19 subjects from day 7 to 14, reaching statistical significance only for CD154+IFN-γ + cells, and did not recover even after the second administration of the vaccine (Figure 3B, C, D and E). However, as observed for B cells, also T cells showed a significant decrease following the second jab. On the contrary, in naïve subjects there was a significant increase in the frequency of SARS-CoV-2 spike-specific CD4+ T cells after the second dose, reaching the frequencies observed in ex COVID-19 subjects. (Figure 3 B, C, D and E). We then looked at the polyfunctional capability of Spike specific CD4+ T cells in naïve and ex COVID-19 subjects at the end of the vaccination protocol, day 28 after the first injection. As shown in Figure 3F, we observed similar frequencies of cells producing 2 or 3 cytokines in combination. In agreement with these findings, we observed that Spike-specific CD4+ T cells displayed a similar expression pattern of two immune checkpoint molecules, TIGIT and PD1 (Figure 3G). Altogether, these data demonstrate that Spike specific CD4+ T cells, at least those identified based on CD154 and cytokine expression, at the end of the vaccination protocol have a similar functional potential in naïve and ex COVID-19 subjects, independently of their prior exposure to SARS-CoV-2.

**Figure 3.**
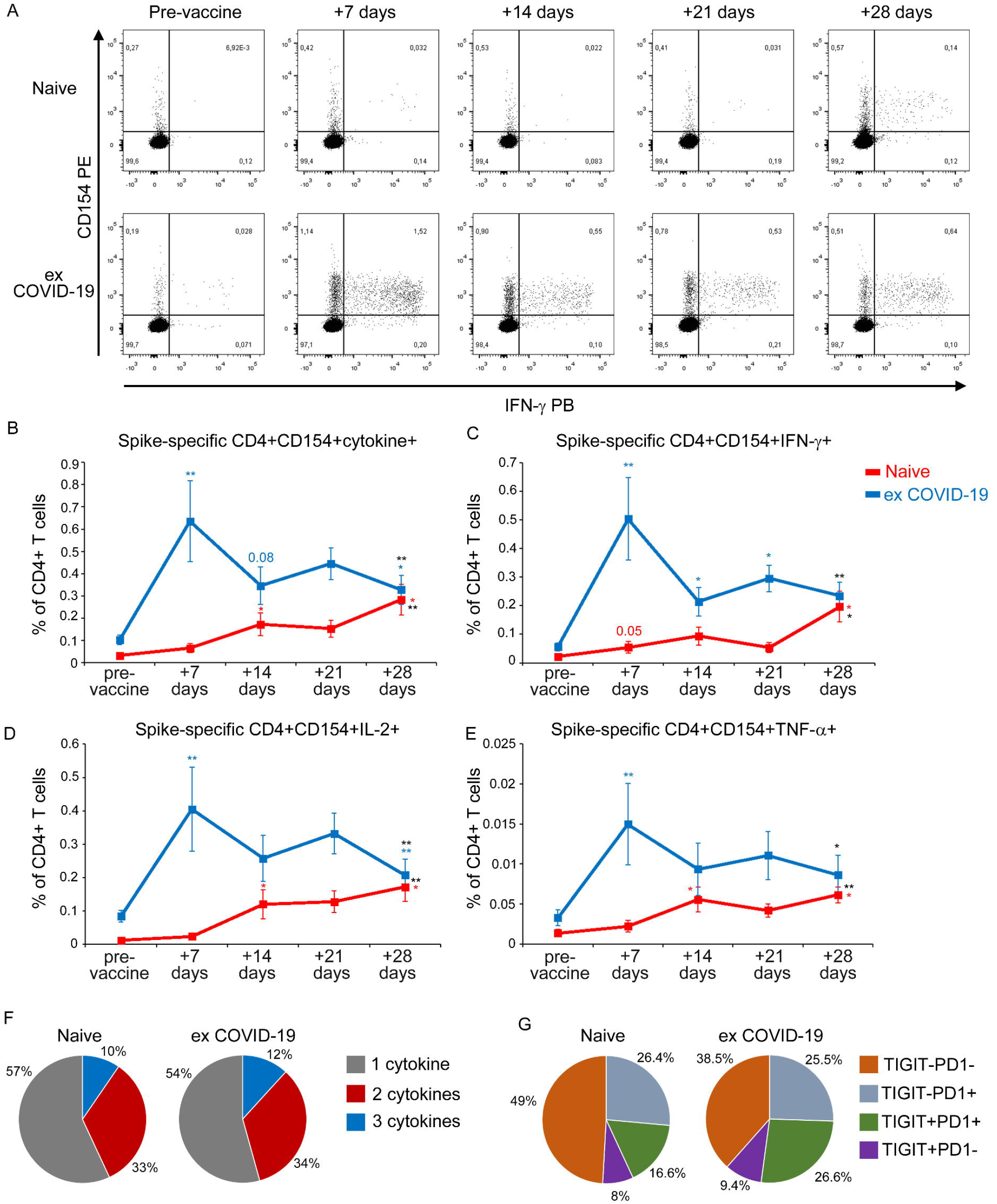
Detection of Spike-specific circulating CD4+ T cells in naïve and ex COVID-19 vaccinated subjects. A) Representative flow cytometric plots of Spike-specific CD4+ CD154+ IFN-γ+ T cells in one naïve (upper row) and one ex COVID-19 (lower row) subject before vaccination and after 7, 14, 21, 28 days from the first injection. Kinetic analysis of frequencies of CD154+ cells producing at least one cytokine among IL-2, IFN-γ and TNF-α (B), CD154+ IFN-γ+ (C), CD154+ IL-2+ (D), CD154+ TNF-α+ (E) Spike-specific T cells in naïve (red lines) and ex COVID-19 (blue lines) subjects, before and after vaccination. Data are represented as mean ±SE from 11 naïve and 11 ex COVID-19 subjects, subtracted of background unstimulated negative control. Blue and red asterisks refer to paired statistics within each study group compared to the previous time point in the kinetic. Black asterisks at day 28 represent paired statistic compared to pre-vaccine point. *= p<0.05; **= p<0.01; ***= p<0.001. (F) Characterization of SARS-CoV-2 Spike-specific CD4+ T cell polyfunctionality in naïve and ex COVID-19 subjects at day 28 following the first vaccine injection. Results are expressed as mean percentages from 11 naïve and 11 ex COVID-19 subjects. (G) Characterization of TIGIT and PD1 expression by SARS-CoV-2 Spike-specific CD4+ T cells in 7 naïve and 8 ex COVID-19 subjects. Results are expressed as mean percentages.

This is the first study, to our knowledge, evaluating the early kinetics of cellular immune response following two doses of BNT162b2 mRNA vaccine in naïve and ex COVID-19 subjects. During the three weeks following the first vaccine injection antibodies, B and T cells specific for SARS-CoV-2 spike protein progressively increase in naïve subjects. In addition to what has been already demonstrated for antibodies and T cells in preclinical studies [11], also B cells increase in the circulation after the second dose. Neutralizing antibody titers remain low, close to cut-off values, until day 21, but maximally increase post second injection, confirming that the second injection is mandatory in naïve individuals [12]. Regarding ex COVID-19 subjects, our findings suggest that there is a rapid reactivation of both humoral and cellular immunological memory to SARS-CoV-2 spike protein. Indeed, 7 days after the first injection we observed maximal increase in circulating spike-specific antibodies, B, and T cells. Interestingly, we observed a decrease in the frequency of SARS-CoV-2 spike-specific CD4+ T cells in ex COVID19 subjects at 14 days after the administration of first vaccine dose. We hypothesize that this reduction could be essentially linked to two phenomena: 1) sequestration of specific CD4+ T cells in the lymph node; 2) suppression of specific CD4+ T cells through tolerogenic mechanisms. It should be noted that B cells do not display the same behaviour, given that their frequency progressively increases until day 21. The administration of second jab instead seems to be ineffective, and it is rather associated to a contraction of both spike-specific circulating B and T cells. These data have been obtained on a relatively small cohort of ex COVID-19 subjects but are in agreement with those presented in the accompanying article by the group of Maria Rescigno, who confirms in a larger cohort that in ex COVID-19 subjects one dose of BNT162b2 is sufficient to maximize spike-specific antibody titers. This information has practical fundamental implications. Indeed, in most countries individuals who recovered from SARS-CoV-2 infection are not excluded from COVID-19 vaccination, and they commonly follow two-doses vaccination schedule [13-14]. However, differently from naïve individuals, these data suggest that one single injection may be sufficient to protect ex COVID-19 subjects in accordance with emerging data [15-16]. We also cannot exclude that the second jab might even be detrimental in this context, leading to a functional exhaustion of spike-specific lymphocytes [17]. Indeed, we observed a decrease in the frequency of both B and T cells at day 28, one week after second dose. Our data demonstrate that spike-reactive CD4+ T cells have a similar polyfunctional capability and immune checkpoint expression pattern in naïve versus ex COVID-19 subjects. However, we cannot exclude the presence of exhausted spike-specific CD4+ T cells that cannot be reactivated ex vivo. It should be noted that our ex COVID-19 cohort includes subjects with a history of symptomatic COVID-19 infection. It is known that the strength of the immune response to SARS-CoV-2 directly correlates to disease severity both in the acute phase [18-19] and in the memory phase [Mazzoni et al., under review] and that memory immune response to SARS-CoV-2 is quite heterogeneous in the months after recovery [20]. For this reason, additional studies are required to understand if one single vaccine administration may be sufficient also in people with a history of asymptomatic SARS-CoV-2 infection or who are characterized by low levels of both humoral or cell mediated antigen-specific immune response. Although approved by international and national medicine agencies, the distribution and administration of vaccines is still limited to a minority of the population. Saving one vaccine dose for each subject who recovered from COVID-19 would substantially increase the number of doses available for naïve individuals, thus shortening the time to reach herd immunity.

## Materials and Methods

### Patients

22 healthcare workers who received BNT162b2 mRNA COVID-19 vaccine, of which 11 had a previous history of SARS-CoV-2 infection (ex-COVID-19), while the others without prior infection, were recruited at the Careggi University Hospital, Florence, Italy, by the Infectious and Tropical Diseases Unit (Supplementary Table S1). Following immunization schedule approved by the European Medicines Agency, each subject received two vaccine injections, 21 days apart. Blood samples were collected before the first dose (basal time, T0) and every 7 days (T7, T14, T21) until T28 (28 days after the first dose, 7 days after the second one) (Supplementary Figure S1).

PBMNCs were obtained following density gradient centrifugation of blood samples using Lymphoprep (Axis Shield Poc As™) and were frozen in FCS plus 10% DMSO to be stored in liquid nitrogen. For each subject, longitudinal samples were defrost and analyzed together for B cells and T cells evaluation. Serum was frozen and stored for Ig levels evaluation.

### Evaluation of SARS-CoV-2-spike-reactive T cells

For T cells stimulation in vitro, 1.5 million PBMNCs were cultured in complete RPMI plus 5% human AB serum in 96 well flat bottom plates in presence of medium alone (background, negative control) or of a pool of spike SARS-CoV-2 peptide pools (Prot_S1, Prot_S+ and Prot_S to achieve a complete sequence coverage of the spike protein) at 0.6 µM/peptide, accordingly to manufacturer’s instructions (Miltenyi Biotech). Staphylococcal enterotoxin B SEB 1 µg/ml (Sigma Aldrich) was used as a positive control. After 2 hours of incubation at 37°C, 5% CO2, Brefeldin A (5 µg/mL) was added, followed by additional 4 hours incubation. Finally, cells were fixed and stained using fluorochrome-conjugated antibodies listed in Table S2 and Table S3. Samples were acquired on a BD LSR II flow cytometer (BD Biosciences). Flow cytometry experiments were performed using published guidelines [21].

### Evaluation of SARS-CoV-2-spike-specific IgM, IgA, IgG

For B cells evaluation, 2 million PBMNCs were stained for 30 minutes at 4°C with fluorochrome-conjugated antibodies listed in Table S4, then washed with PBS/EDTA buffer (PEB), and incubated 5 minutes with 7-AAD for viability evaluation. Samples were acquired on a BD LSR II flow cytometer (BD Biosciences). Recombinant biotinylated SARS-Cov2 spike protein (Miltenyi Biotech) was conjugated separately with streptavidin PE and PE-Cy7 for 15 minutes at room temperature and pooled in 1:2 ratio, before being added to final staining mix.

### Evaluation of SARS-CoV-2-specific IgM and IgG

Evaluation of SARS-CoV-2-spike specific IgG (Diasorin) and IgM (Abbott), Nucleoprotein-specific IgG (Abbott), RBD-specific IgG (Abbott), S-neutralizing Ig (Dia.Pro Diagnostic Bioprobes) were performed following manufacturer’s instructions. The antibody reactivity of each specimen was expressed by the ratio between optical density and cut-off value (index) or as arbitrary units/ml.

### Statistics

Unpaired Mann-Whitney test was used to compare ex COVID-19 subjects versus naïve subjects; Wilcoxon-Signed Rank test was used to compare different time points in each study group. In all cases, p values ≤0.05 were considered significant.

### Study approval

The procedures followed in the study were approved by the Careggi University Hospital Ethical Committee. Written informed consent was obtained from recruited patients.

## Data Availability

All the relevant data are included in the manuscript.

## Acknowledgements

We thank all the subjects who participated to the study. This study was supported by funds to the Department of Experimental and Clinical Medicine, University of Florence, derived from Ministero dell’Istruzione, dell’Università e della Ricerca (Italy) (Project Excellence Departments 2018-2022) and by University of Florence, project RICTD2122.

## COVID-19 Research Group

Silvia Benemei, Irene Campolmi, Alberto Farese, Filippo Lagi, Marco Matucci Cerinic, Fabrizio Niccolini, Diana Paolini, Marco Pozzi, Matteo Tomaiuolo

## Author Contributions

A.M., N.D.L, F.L., G.M.R., L.C., A.B., F.A., designed the study; N.D.L., E.M., M.S., L.Z. collected peripheral blood samples and collected informed consent; A.M., L.M., L.S., A.V., M.C., G.L., S.T.K., M.G.C., performed experiments; P.P., C.S, L.T., provided advice; A.M., L.M., analyzed data; A.M., L.S., F.A. wrote the manuscript. All authors revised the manuscript and gave final approval.

## Notes

### Competing Interest Statement

The authors have declared no competing interest.

### Author Declarations

This work has been approved by Careggi ethical committe (Ref. 16859; 19466)

