## Supplementary data for "First dose mRNA vaccination is sufficient to reactivate immunological memory to SARS-CoV-2 in ex COVID-19 subjects"

**Supplementary Table S1. Demographic and clinical features of enrolled subjects.**

| Case ID | Gender | Age at first dose vaccination | COVID-19 | Time from COVID-19 resolution to first dose vaccination (months) |
|---|---|---|---|---|
| **Ex-COVID-19** | | | | |
| **EC1** | F | 51 | severe | 7 |
| **EC2** | F | 52 | mild | 7 |
| **EC3** | M | 60 | severe | 8.5 |
| **EC4** | M | 65 | moderate | 9 |
| **EC5** | F | 53 | moderate | 8 |
| **EC6** | M | 37 | moderate | 8.5 |
| **EC7** | F | 54 | mild | 8 |
| **EC8** | M | 61 | critical | 9 |
| **EC9** | M | 64 | critical | 9 |
| **EC10** | F | 51 | mild | 6 |
| **EC11** | F | 44 | moderate | 8 |
| **Mean (years)** | - | 53,8 | - | - |
| **SD (± years)** | - | 8,5 | - | - |
| **Non-COVID-19** | | | | |
| **NC1** | F | 35 | no | na |
| **NC2** | F | 61 | no | na |
| **NC3** | F | 44 | no | na |
| **NC4** | F | 41 | no | na |
| **NC5** | M | 41 | no | na |
| **NC6** | M | 32 | no | na |
| **NC7** | F | 37 | no | na |
| **NC8** | F | 29 | no | na |
| **NC9** | F | 46 | no | na |
| **NC10** | F | 24 | no | na |
| **NC11** | M | 53 | no | na |
| **Mean (years)** | - | 40,3 | - | - |
| **SD (± years)** | - | 10,7 | - | - |

*na denotes not applicable*

**Supplementary Table S2. List of all fluorochrome mAbs used for flow cytometric analysis of antigen specific T cells.**

| Antigen | Flurochrome | Clone | Company |
|---|---|---|---|
| TNF-α | FITC | 6401.1111 | BDBioscience |
| CD154 | PE | TRAP1 | BDBioscience |
| CD3 | PerCP | SK7 | BDBioscience |
| CD4 | PE-Cy7 | SK3 | Invitrogen |
| CD8 | Super Bright 600 | SK1 | eBioscience™ |
| IL-2 | APC | MQ1-17H12 | BDBioscience |
| IFN-γ | Pacific Blue | B27 | BioLegend |
| L/D | Fixable Viability Stain 780 | | BDBioscience |

**Supplementary Table S3. List of all fluorochrome mAbs used for flow cytometric analysis of immune checkpoint expression by antigen specific T cells.**

| Antigen | Flurochrome | Clone | Company |
|---|---|---|---|
| TNF-α | FITC | MAb11 | BDBioscience |
| IFN-γ | FITC | 25723.11 | BDBioscience |
| IL-2 | FITC | 5344.111 | BDBioscience |
| CD8 | PerCP | SK1 | BDBioscience |
| CD4 | eFluor506 | RTA-T4 | Invitrogen |
| CD3 | Pacific Blue | UCHT1 | BDBioscience |
| PD1 (CD279) | PE-Cy7 | EH12.2H7 | BioLegend |
| TIGIT | APC | MBSA43 | Invitrogen |
| CD154 | PE | TRAP1 | BDBioscience |
| L/D | Fixable Viability Stain 780 | | BDBioscience |

**Supplementary Table S4. List of all fluorochrome mAbs used for flow cytometric analysis of antigen specific B cells.**

| Antigen | Flurochrome | Clone | Company |
|---|---|---|---|
| CD19 | APC-Vio770 | LT19 | Miltenyi |
| CD27 | VioBright FITC | M-T271 | Miltenyi |
| IgA | VioGreen | IS11-8E10 | Miltenyi |
| IgG | VioBlue | IS11-3B2.2.3 | Miltenyi |
| CD14 | PerCP | TÜK4 | Miltenyi |
| IgM | APC | PJ2-22H3 | Miltenyi |
| CD3 | PerCP | BW264/56 | Miltenyi |
| 7AAD | | | Miltenyi |
| Spike | PE | | Miltenyi |
| Spike | PE-Vio770 | | Miltenyi |

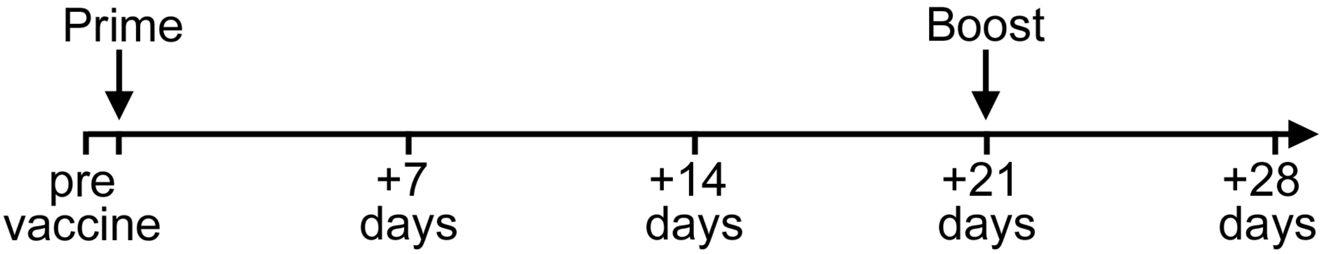

**Supplementary Figure S1. Vaccination schedule and time points of analysis**

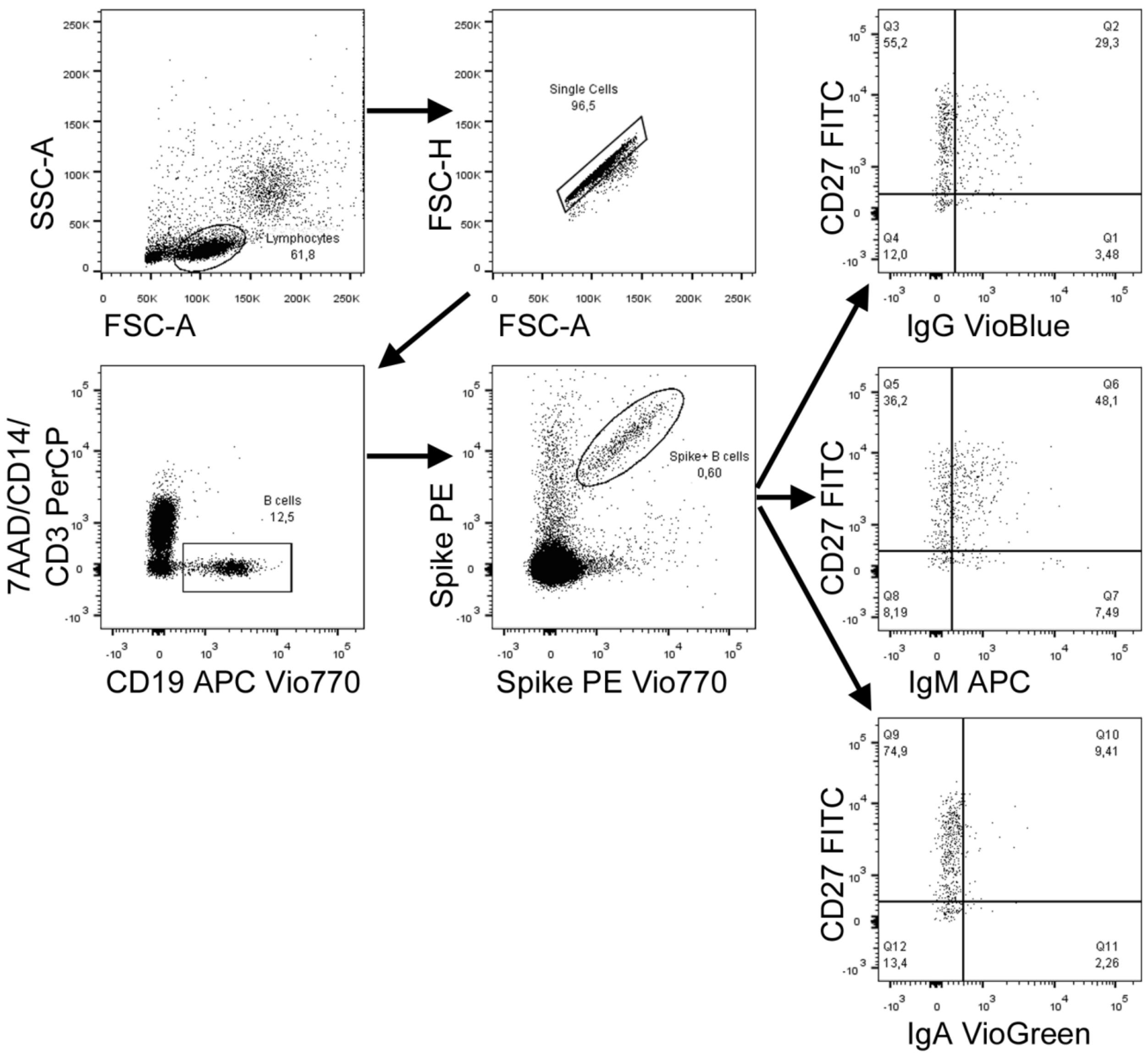

**Supplementary Figure 2: Gating strategy for the identification of spike-specific B cells**

Lymphocytes were gated based on physical parameters (FSC-SSC), then doublets were removed using FSC-A and FSC-A parameters. PerCP was used as dump channel for the exclusion of dead cells (7AAD), CD3+ T cells and CD14 monocytes. B cells were identified as CD19+. B cells binding PE- and PE Vio770-conjugated spike protein were then identified as spike-specific. Among spike-specific B cells we evaluated CD27 expression associated to IgG, IgM and IgA.

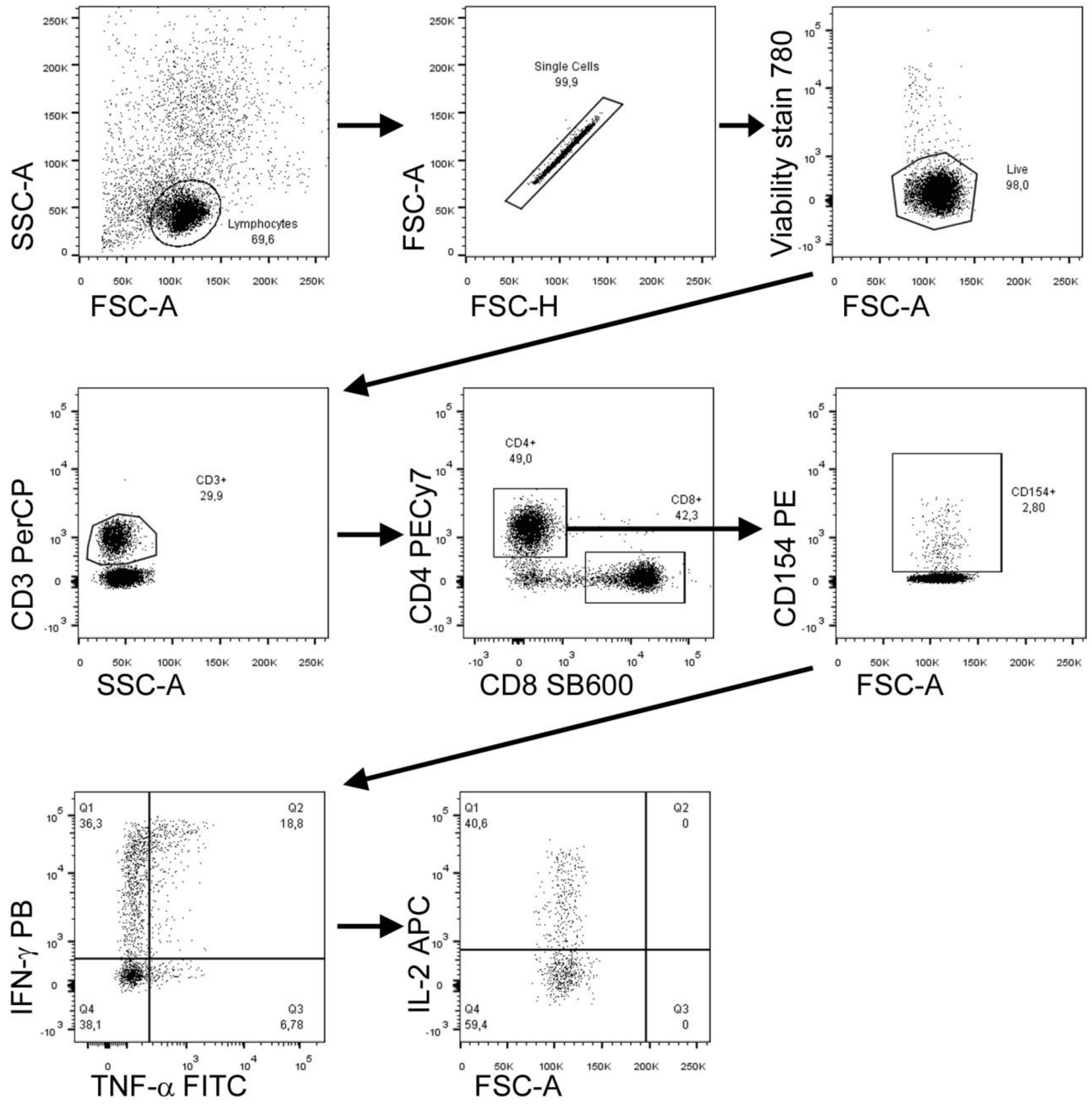

**Supplementary Figure 3: Gating strategy for the identification of spike-specific T cells**

Lymphocytes were gated based on physical parameters (FSC-SSC), then doublets were removed using FSC-A and FSC-A parameters. Dead cells were excluded using viability stain 780. T cells were identified as CD3+. We then identified CD4+ T cells. Among these, we identified CD154 expressing cells. CD4+CD154+ T cells were then evaluated for IFN-γ, TNF-α and IL-2 expression.
